# A systematic review of the factors associated with suicide attempts among sexual-minority adolescents

**DOI:** 10.1101/2022.01.19.22269164

**Authors:** Xavier Wang, Quan Gan, Junwen Zhou, Mireille Cosquer, Bruno Falissard, Emmanuelle Corruble, Catherine Jousselme, Florence Gressier

## Abstract

**Introduction:** Suicide attempt is a worldwide major public health problem, that accounts for 1.4% of all deaths worldwide Recent literature reported higher risk of suicide attempt among adolescents associated with sexual minority status but few systematic reviews focused on the risk and protective factors. For that reason, it seems necessary to examine risk and protective factors of attempted suicide in sexual minority adolescents.

**Methods:** We conducted a systematic review of published studies on factors associated with suicide attempts in LGBT adolescents. 4 databases up to December 2020 were searched to find relevant studies.

**Results:** In addition to the factors usually found in general population (gender, ethnic minorities, childhood trauma, psychiatric symptoms, addictive behaviors), some factors have been independently associated with suicide attempt in LGBT population: early coming out, not acceptable by families, not satisfied with LGBT friendship, too few number of friends, physical abuse, sexual abuse and bullying. The protective factors of suicide attempt reported in LGBT population were feeling safety at school, teacher support, anti-bullying policy, and other adult’s support.

**Conclusion:** Effective preventive measures for suicide attempt among LGBT youth need to be developed and implemented. The impact of interventions targeting teacher responses to LGBT stigma, discussion of LGBT issues in class and reactions of family and friends to the coming out of LGBT youth should be further investigated.

## Background

Mental health conditions account for 16% of the global burden of disease and injury in people aged 10-19 years and suicide is the fourth leading cause of death in 15-19-year-olds (World Health Organisation, 2020). Among all the mental health problems in adolescents, suicide attempt is a worldwide major public health problem, that accounts for 1.4% of all deaths worldwide (Smith et al., 2020). In the United States, it has become the second leading cause of death among youth aged 10-24 since 2011, which caused the loss of 6000 lives a year in the year’s 2011 to 2019 (Centers for Disease Control and Prevention, 2019). In addition, the situation of suicide attempt among adolescents associated with sexual minority status is more severe (Greteman, 2015; Meyer, 2003; Ueno, 2005; Williams et al., 2021). Lesbian, gay and bisexual (LGB) adolescents are 2 to 8 times more likely to have attempted suicide compared to their heterosexual peers (Firdion & Beck, 2015; Huang et al., 2018; Needham & Austin, 2010; Seil, Desai, & Smith, 2014; Williams et al., 2021). Some studies in the United States showed that adolescents between 15-19 years of age with a homosexual attraction experience more SA than their heterosexual peers (Seil et al., 2014; Teasdale & Bradley-Engen, 2010). In Europe, a study in Iceland looked at the problem of the risk of SA among LGB adolescents between 15 and 16 years old. It reported that the risk of SA was 4 to 6 times higher in LGB than in heterosexual adolescents (Arnarsson, Sveinbjornsdottir, Thorsteinsson, & Bjarnason, 2015).

Due to the growing frequency for suicide attempt during the last decades, adolescents are the main target population of prevention policies to reduce suicide and the identification of suicide risk factors is crucial. Many studies have reported factors including mood disorders (depression and anxiety), substance abuse and prior suicide attempts are strongly related with youth suicides (Pelkonen & Marttunen, 2003; Swee, Shochet, Cockshaw, & Hides, 2020). Anxiety and feelings of pressure linked in large part to fear of the judgment of others, often refrain LGB adolescents from sharing their sexual orientations, which seems to push them more frequently than their heterosexual peers to have suicide attempts (Hatzenbuehler, 2011; Liu & Mustanski, 2012). Factors related to family adversity, social exclusion and poor school performance also contribute to the risk of suicide (Hernández-Bello, Hueso-Montoro, Gómez-Urquiza, & Cogollo-Milanés, 2020; Pelkonen & Marttunen, 2003). Numbers of risks have made adolescents vulnerable to negative results, but protective factors have facilitated a resilient response, so both protective and risk factors should be considered in terms of factors that may link to suicide attempts among adolescents (Joiner et al., 2009). According to the protective environments, Bronfenbrenner’s ecological systems theory (Hong, Espelage, & Kral, 2011) has divided the protective factors of suicide into several systems levels (micro, meso, exo, macro and chrono). Because different environments will act on different factors which will produce different effects. Moreover, as a sexual minority group, due to their sexual orientation, LGBTs may even experience more negative factors than heterosexual people, leading to a further increase in suicide risk (Williams et al., 2021). One study in UK revealed that psychological factors may account for the association between LGB status and suicide attempt (Taylor, Dhingra, Dickson, & McDermott, 2020). During the last decades, different experts have reported systematic review of prevalence rather than risk or protective factors for suicide attempts in LGBT adolescents (Miranda-Mendizábal et al., 2017; Pelkonen & Marttunen, 2003; Williams et al., 2021). Considering the significant risk of suicide among LGB youth, it seems essential to identify the risk and protective factors.

### Objective

Our aim is to systematically review the literature on the risk and protective factors of suicide attempt in sexual minority adolescents.

## Method

### Eligibility Criteria

#### Inclusion criteria

a. The target population is sexual minority teenagers with definition of sexual orientation (gay, lesbian, homosexual, bisexual) and/or reported levels of same sexual attraction or behavior;
b. The age includes 13-20 years’ old (World Health Organisation, 2020)
c. With clearly identified suicide attempt as outcome;
d. With definite risk or protective factors of suicide attempt in the result
e. all types of studies: cross-sectional studies, cohort studies and case-control studies.
f. No language limitation.

### Information sources and Search

Two authors (X.W and Q.G) searched four databases (PubMed, Web of science, Cochrane library and PsycInfo) for articles published until 31 December 2020 using the search strategy “suicide” and “adolescent” or “youth” or “young people” or “teenager” and “LGB” or “gay” or “lesbian” or “homosexual” or “sexual minority” or “sexual orientation”. The disagreements were resolved by discussion.

### Study selection

From 422 articles identified, 60 duplicates were removed. After screening the titles and abstracts, 281 were excluded for the following reasons: 125 did not concern suicide attempt; 86 did not describe any risk or protective factor of suicide attempt; 47 did not include the participants aged 13-20 years old; 17 were based on a population not including sexual minorities; 6 only concerned transgender population. After reviewing the full text papers, 69 were excluded for the following reasons: 50 did not describe any risk or protective factor of suicide attempt; 10 were based on second analysis of published articles; 6 did not include the youth participants; 2 did not concern suicide attempt; 1 only concerned transgender population. Finally, 12 articles met the inclusion criteria (Flow chart 1).

**Flow chart 1.**
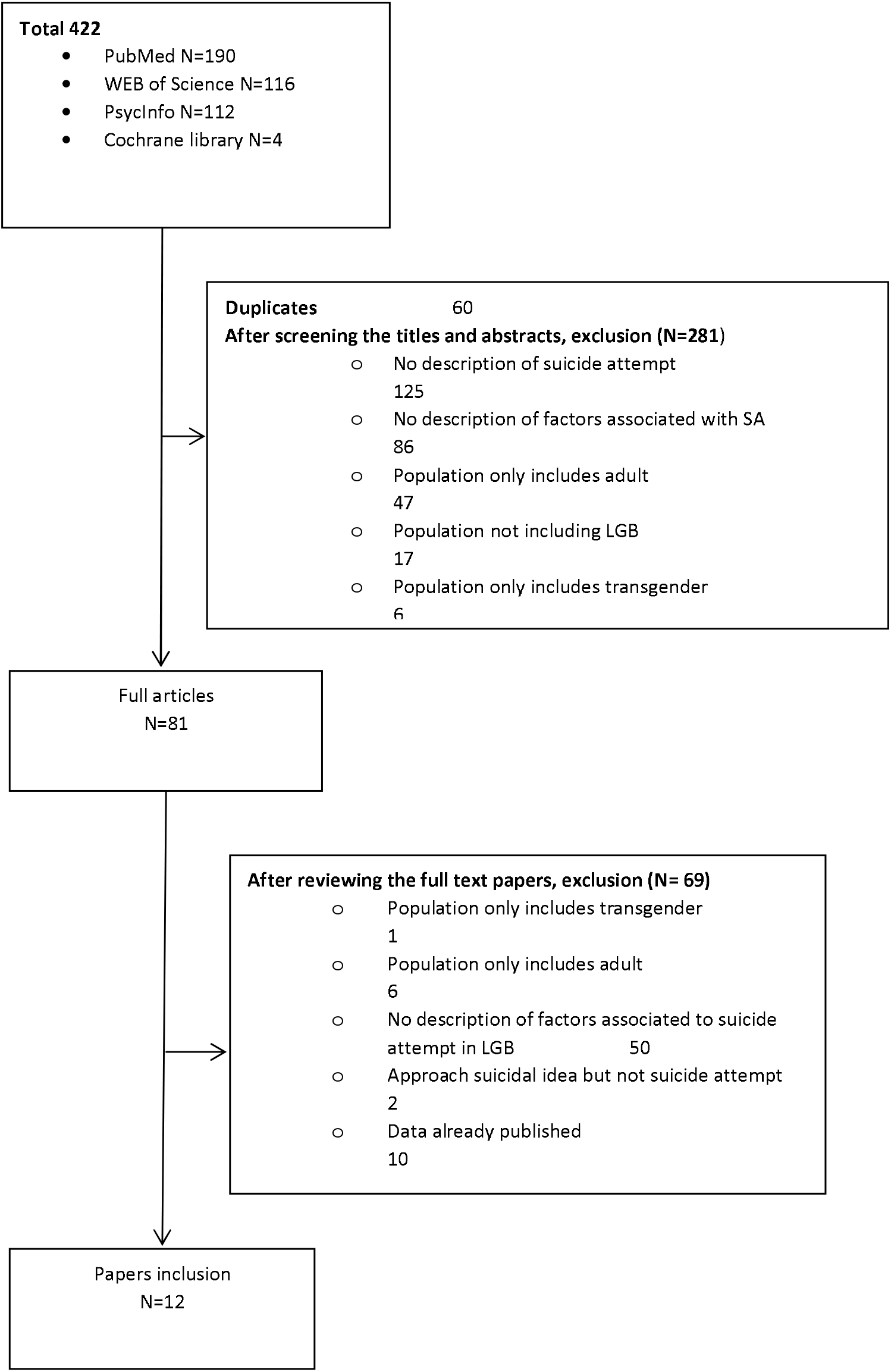

### Data collection process

The titles and abstracts were screened independently by two authors (X.W. and Q.G.). After selecting the efficient articles, they have been downloaded and read. One author (X.W) extracted the data and entered in the forms. Two authors (C.P and Q.G.) examined and verified the information entered was properly.

The following information was extracted in table 1: Author, publication year, country, target population, age range, study design, total sample size, sexual minority sample size, definition and proportion of suicide attempt, quality assessment.

**Table 1.** Characteristics of included articles

| Author | Country | Population | Age | Study design | Suicide attempt | Total sample size | LGB size (% girls) | Proportion of suicide attempts LGB | Quality of study |
| --- | --- | --- | --- | --- | --- | --- | --- | --- | --- |
| Humphries et al. 2020 | US | A nationally representative sample of students in grades 9<sup>th</sup> to 12<sup>th</sup> enrolled in either public or private schools | 14-18 | cross-sectional | last 12 months | 27706 | 2740 | 25.8% | 9 |
| Busby et al. 2020 | US | 868 students from four universities who completed an online screening survey | 18-30 | cross-sectional | lifetime | 868 | 868 (63.6%) | 23% | 8 |
| Turpin et al. 2020 | US | all regular public and private schools with students in at least one of 9th to 12th grades in the 50 states and the District of Columbia | 12-18 | cross-sectional | last 12 months | 876 | 876 | 29.5% | 9 |
| Toomey et al. 2019 | US | a large sample of in-school US adolescents | 11-19 | cross-sectional | lifetime | 116925 | 5598 | 37.7% | 6 |
| Rimes et al. 2019 | UK | LGB young adults | 16-25 | cross-sectional | lifetime | 3275 | 3275 (49.3%) | 13.6% | 9 |
| McDermott et al. 2017 | UK | community-based via LGBT organizations and social media (twitter, FB, Tumblr) | 13-25 | mixed study | lifetime | 789 | 789 (42.6%) | 17.6% | 4 |
| Taliaferro et al. 2017 | US | population-based survey administered every 3 years to students in grades 5, 8, 9, and 11 | 14-17 | cross-sectional | last 12 months | 77758 | 2878 | 12.5% (Gay or les), 19.5% (Bisexual) | 9 |
| Duong et al. 2014 | US | the 11,877 students enrolled in grades 9 through 12 from 105 NYC public high schools | 14-18 | cross-sectional | last 12 months | 11877 | 951 (69.5%) | 27.3% | 9 |
| Hatzenbuehler et al. 2013 | US | random sampled from 11th grade school students | 13-17 | cross-sectional | last 12 months | 31852 | 1413 (67.4%) | 21% LG, 23% B | 8 |
| Mustanski et al. 2013 | US | venue sampling (flyers in neighborhoods by LGBT youth 38%) and snowball sampling (incentivized recruitment of peers by existing participants 62%) | 16-20 | cross-sectional | whole life, last year | 237 | 237 (52.3%) | 31.6% (lifetime) 7.2% (last year) | 10 |
| Goodenow et al. 2006 | US | a population-based survey of adolescents from 64 public high schools | 14-18 | cross-sectional | last 12 months | 3607 | 202 (49%) | 28.5% | 10 |
| Heerngen et al. 2000 | Belgium | a general population sample of homosexual or bisexual young people and a control sample consisting of secondary and high school students | 15-27 | cross-sectional | lifetime | 404 | 219 (37.4%) | 17.2% | 9 |

### Data synthesis

We extracted odds ratio and 95% confidence intervals of risk and protective factors from the results of multiple logistic regressions that focused on LGBT participants in 12 articles. We also divided all influencing factors into three categories according to the environmental contexts: personal environment (demographic, traumatic state, health condition, consumption of substances, psychiatric disorder, personal life), intimate environment (family/adult support, friendship) and public environment (school, internet, society).

The following information was extracted in table 2: Year of study, name of investigation, confirmed risk or protective factors with odds ratio and 95% confidence intervals, non-significant risk or protective factors with odds ratio and 95% confidence intervals, adjustment variables and conclusion.

**Table 2.**
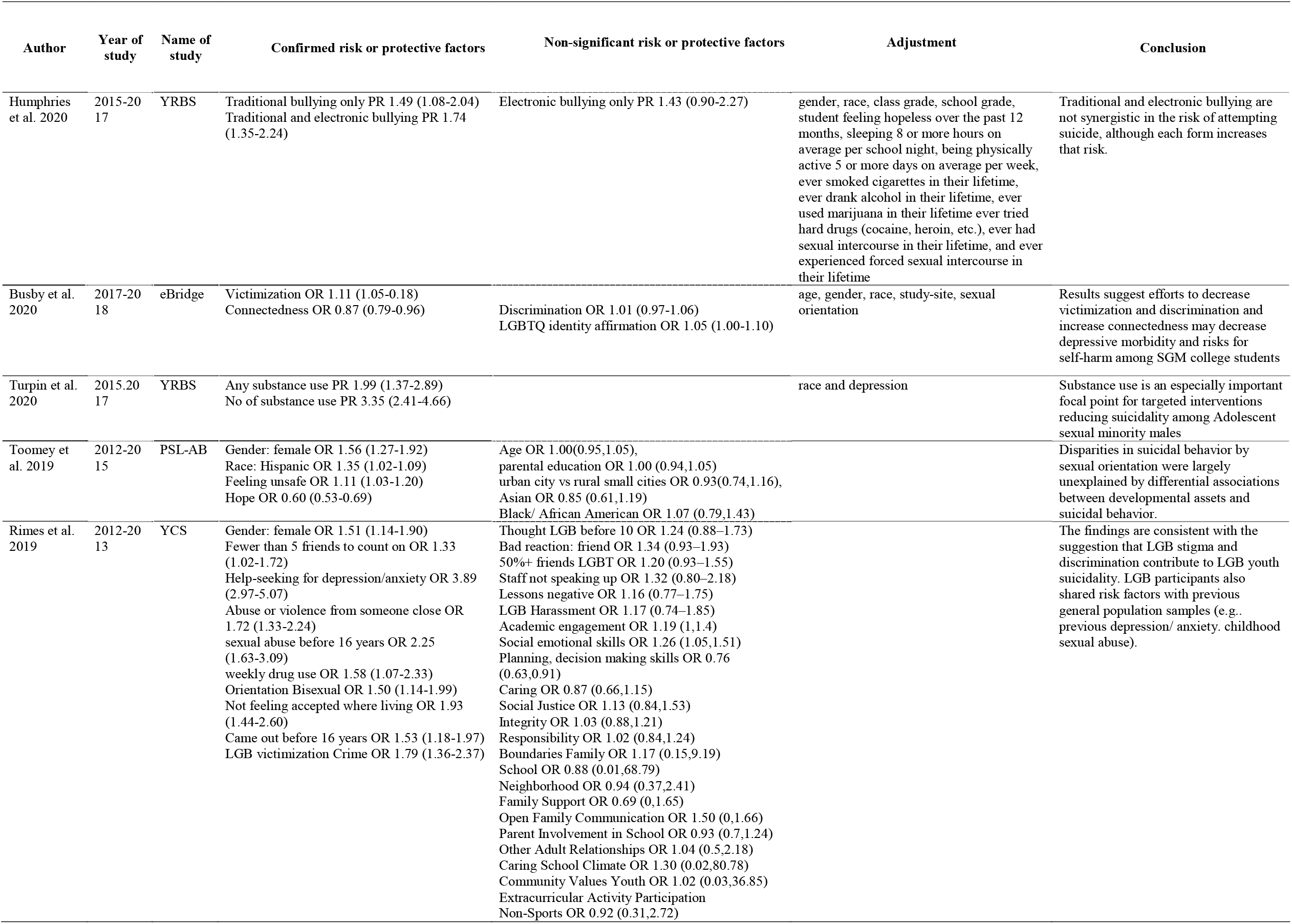
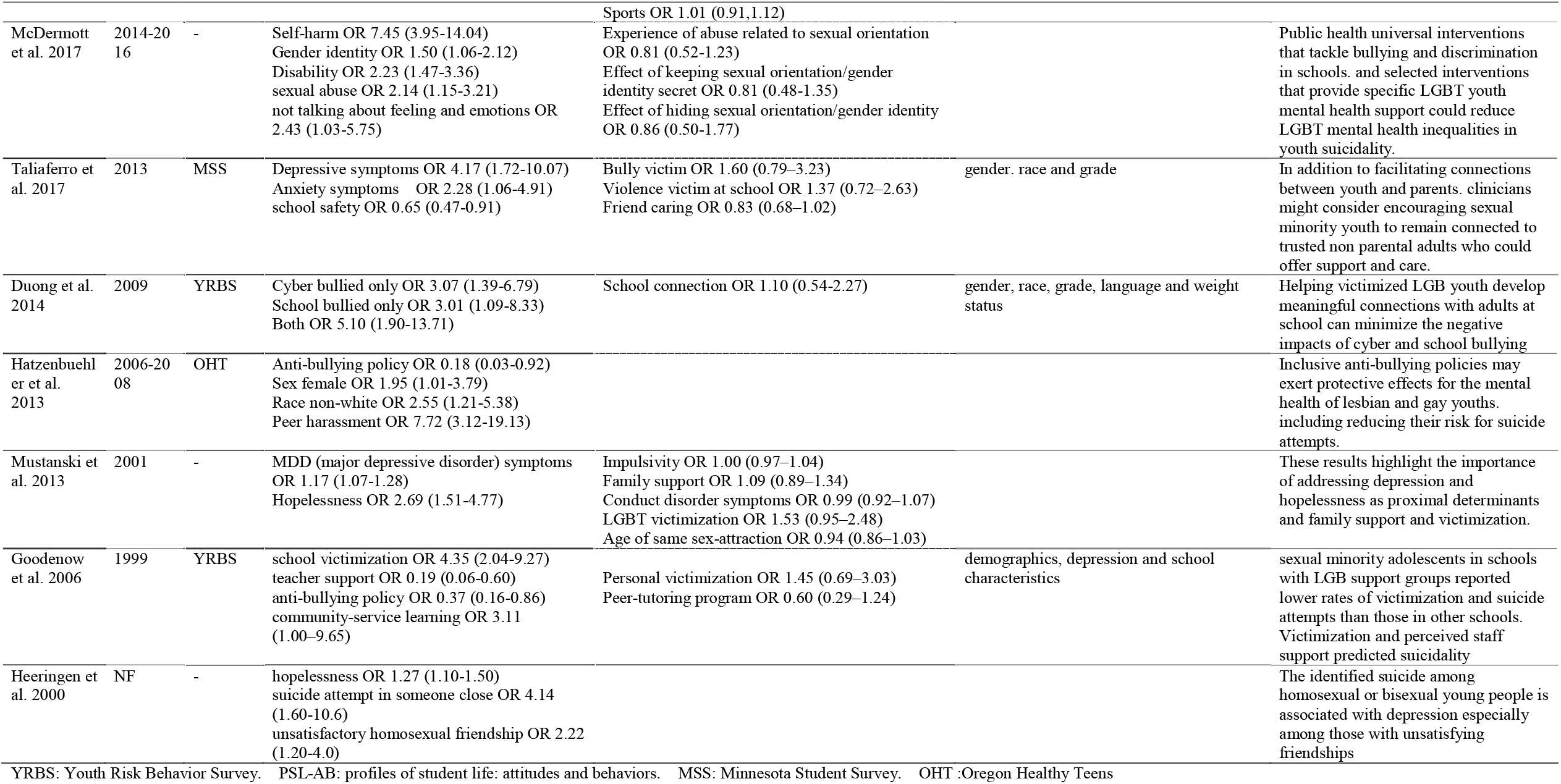
information of risk and protective factors in LGB adolescents

### Quality assessment of the studies

The Newcastle-Ottawa scale (Stang, 2010) was used to evaluate the quality of include studies. This scale is widely used as evaluation tool of observational studies and longitudinal studies (Moskalewicz & Oremus, 2020). It has three categories including eight entries with a full score of 10. We classify 8-10 points as high quality, 5-7 points as medium quality, less than 5 points as low quality (Appendices).

## Result

### Study characteristics

Of the 12 articles included, 10 studies were identified as high quality, 1 was moderate quality and only 1 was low quality. All but one was cross-sectional studies. 9 studies were based in the US, 2 in the United Kingdom, 1 in Belgium.

### Population

6 studies involved the subjects aged between 12 and 18 years old, 2 until 20 years, 2 until 25 years, and 2 studies involved people aged until 30 years. All studies included at least two genders. 5 were on LGBT only. 7 took into account both sexual minority and heterosexual.

### Definition of sexuality

Of 12 studies, 8 studies used one question to define sexual orientation, of which 4 studies originated from the same project YRBS (Youth Risk Behavior Survey) in U.S., but focused on different regions or years. The question they used was “Which of the following best describes you?” The answers included: heterosexual (straight); gay or lesbian; bisexual; not sure. 2 studies used sex behavior by asking the question “With whom have you had sexual contact?” Response options included no one, female(s), male(s), both female(s) and male(s). And 2 studies did not mention the definition.

### Definition of suicide attempts

6 studies collected the information of attempt suicide during last 12 months by the question “During the past 12 months, how many times did you actually attempt suicide?” In 5 studies, participants described whether they ever had attempted suicide in their life. 1 study collected the data of both whole life and last year.

### Associated factors

#### Personal environment

Considering personal environment, we analyzed risk factors associated with suicide attempt in adolescents and also risk factors more specifically related to sexuality.

#### Demographic

We categorized 5 elements (gender, race, age, rurality, parental education) in demographic category. Gender and race were considered as definite risk factors in sexual minority adolescents. Age, rurality and parental education were analyzed only in one study reporting no difference (Toomey, Syvertsen, & Flores, 2019). For gender, 4 studies reported that girls were more likely to commit suicide than boys (Hatzenbuehler & Keyes, 2013; McDermott, Hughes, & Rawlings, 2018; Rimes et al., 2019; Toomey et al., 2019). Three studies adjusted their results for gender (Taliaferro, et al., 2017; Duong et al., 2014; Goodenow et al., 2006). For ethnicity, one study has revealed that non-white participants had higher risk of suicide attempt than white participants (Hatzenbuehler & Keyes, 2013). And study of Toomey implied that the Hispanic youth had higher risk than other ethnicities (Toomey et al., 2019).

#### Substances Consumption

Only two studies interested in substance abuse (Rimes et al., 2019; Turpin, Rosario, & Dyer, 2020). The consumption of substance was considered as a risk factor in the study of Turpin who reported the number of substances used (0-7) was identified the strongest associations with suicide attempt (Turpin et al., 2020). The study of Rimes revealed the same result that weekly drug use increased the risk of suicide attempt in LGBT youth (Rimes et al., 2019).

#### Psychiatric disorder

Depression or anxiety was evaluated in three studies reporting higher risks of suicide attempt (Mustanski & Liu, 2013; Rimes et al., 2019; Taliaferro & Muehlenkamp, 2017). Hopelessness was studied and found as risk factor in three studies (Toomey et al., 2019; Mustanski and Liu, 2013. Heeringen et al., 2000). Moreover, not talking about feelings and emotions (McDermott et al., 2018) and feeling unsafe (Toomey et al., 2019) were also reported as risk factors. Furthermore, a suicide attempt in someone close was found to be a risk factor (van Heeringen, Vincke, & van Heeringen, 2000).

#### Traumatic state

Physical abuse and sexual abuse were also identified as risk factors (McDermott et al., 2018; Rimes et al., 2019). And childhood abuse or violence from someone close was also associated with suicide attempt (Rimes et al., 2019).

#### Personal Life

Few studies interested in personal sexual life include age of thought LGBT, coming out and experience. The adolescents coming out before 16 years of age were more likely to commit suicide than others (Rimes et al., 2019) while another study revealed no association between LGBT identity affirmation and suicide behavior (Busby et al., 2020). Whereas self-identified as LGBT before the age of 10 was reported no association with suicide attempt (Rimes et al., 2019), and the age of being attracted to the same sex was not associated with suicide behavior (Mustanski & Liu, 2013).

#### Intimate environment

##### Family/adult support

Only 3 studies were interested in family/adult support but providing inconsistent results. Goodenow found that an adult (teacher) support was associated with a protective effect against suicide attempts (Goodenow, Szalacha, & Westheimer, 2006). However, Mustanski did not report such association (Mustanski & Liu, 2013). Rimes has focused in 4 elements of support (family support, open family communication, parent involvement in school and other adult relationships), unfortunately, none of these showed significance in multiple regression model (Rimes et al., 2019). In addition, not feeling accepted where one lives, doubled the risk of suicide attempts (Rimes et al., 2019).

#### Friend

Friend relationship and support were analyzed in 4 studies. Van Heeringen found that if the relationships of LGB peers around them are not satisfactory, the suicide attempt of homosexual adolescents will be greatly increased (van Heeringen et al., 2000). Too few friends (less than 5 friends) can also increase the risk of suicide attempt (Rimes et al., 2019). However, two studies reported no associations with a negative reaction of a friend or most friends have the same sexual orientation (Rimes et al., 2019) as well as friend caring (Taliaferro & Muehlenkamp, 2017). Social connectedness was a protective factor to prevent the suicide attempt (Busby et al., 2020). The protective policy of peer-tutoring program didn’t show any significance in Goodenow’s study (Goodenow et al., 2006).

#### Public environment

##### School

School environment was analyzed in 6 studies. Two studies reported the importance of school safety: From Taliaferro’s study, perceived safety at school was considered as a protective factor protected against a suicide attempt of gay/lesbian youth (Taliaferro & Muehlenkamp, 2017) and feeling unsafe at school in Toomey’s study showed a risk of suicide attempt (Toomey et al., 2019). Whereas the studies of Duong and Rimes did not find any association with school connection and a caring school climate (Duong & Bradshaw, 2014; Rimes et al., 2019). According to school victimization, Goodenow reported an association of suicide attempt with school victimization (Goodenow et al., 2006). However, Taliaferro reported no association with violence victim at school (Taliaferro & Muehlenkamp, 2017). Other elements of school lessons referring negatively to LGBT issues and school staff not consistently speaking up against LGBT prejudice were reported as no significance (Rimes et al., 2019).

##### Internet

Cyber bullying is also considered a strong risk factor of suicide attempt in sexual minority youth. The study of Duong and Bradshaw divided bulling into school bullying and cyber bullying, while suffering from both types of bullying at the same time has the highest risks (Duong & Bradshaw, 2014). However, another study didn’t find association between electronic bullying and suicide attempt, it only reported the significance of traditional bullying or both traditional and electronic bullying (Humphries, Li, Smith, Bridge, & Zhu, 2021).

##### Society

Among all the risk factors, bullying is always considered to be the most important factor (Hatzenbuehler & Keyes, 2013). The bullying that targets sexual orientation was also thought to be a risk factor, one study (Hatzenbuehler & Keyes, 2013) found that the homosexual population had higher risks of being bullied than the bisexual population. Rimes also reported that LGBT victimization crime was as risk factor of suicide attempt (Rimes et al., 2019). However, experience of abuse related to sexual orientation was not associated in one study (McDermott et al., 2018). Therefore, regarding the protective factors, the most important is the anti-bullying policy. One study (Hatzenbuehler & Keyes, 2013) found “this policy was associated with reduced risk for suicide attempts among lesbian and gay youths”, another study (Goodenow et al., 2006) revealed that “anti-bullying policy significantly predicted a lower probability of single or multiple suicide attempts” in LGBT adolescents.

## Discussion

Majority of studies and meta-analyses have focused on the prevalence of suicide attempt rather than investigating risk factors (di Giacomo, Krausz, Colmegna, Aspesi, & Clerici, 2018; Miranda-Mendizábal et al., 2017). Another precedent review focused on investigating risk factors in sexual minorities population rather than adolescents (Yildiz, 2018). The current systematic review analyzed in detail the LGBT youth suicide risk factor and gave specific classifications.

### Personal environment

From the prospective of cognitive molding, risk or protective factors may have a corresponding continuity, from adolescence to adulthood. Corresponding literatures also corroborate such a proposition (Oi & Wilkinson, 2018). According to the cumulative disadvantage perspective, constant exposure to risk, like victimization and marginalization, could be classified as personal environment and public environment that may lead to an increase in the likelihood of a given outcome (Dannefer, 2003). But there are also some reports in the literature that under the implementation of protective policies, significant results were found in the pattern of declining depressive symptoms among LGBT population (Cardom, Rostosky, & Danner, 2013; Everett, 2015).

In addition, ethnic minorities also showed a higher risk of suicide attempt than Caucasian ethnicities (Hatzenbuehler & Keyes, 2013; Toomey et al., 2019), which also implies that ethnic minorities that living in Western countries were in a more vulnerable situation in terms of their sexual orientations. Many factors associated with sexuality have also been studied: Coming out during adolescence at an early age (before 16 years old) is considered to be a risk factor (Rimes et al., 2019), which may generate more family rejections and school bullying (Annor et al., 2018; Humphries et al., 2021; Needham & Austin, 2010; Ryan, Huebner, Diaz, & Sanchez, 2009). And the result will lead to more LGBT victimization in school and emotional or physical blames by family members. This also suggests that coming out before maturity may hinder the determination of sexual orientation, especially in the dual hostile environment with a lack of family support and anti-LGBT attitudes. It was consistent with another study reporting that sexual orientation identity affirmation is no longer a risk factor for suicide attempt in college students over 18 years old (Busby et al., 2020). From this study, we got that after the formation of sexual orientation with the completion of puberty, LGBT young people can face their identity and orientation more calmly.

### Interpersonal environment

Relevant literature still showed that relationship discrimination and low-quality intimate relationships, either family relationships, or friendships, are major risk factors for the adult LGBT community (Yildiz, 2018). On the basis of odds ratios, LGBT young adults who reported higher levels of family rejection during adolescence were 8.4 times more likely to report having attempted suicide (Ryan et al., 2009). The family support is considered to be one of the most important direct environments for the growth of adolescents. Some studies have shown that parental support is more important than peer support (Mustanski & Liu, 2013; Rimes et al., 2019). Therefore, the acceptance of parents to receive education on minority sexual orientations should also be the core of policy formulation.

According to LGBT victimization, sexual minorities will feel tremendous pressure due to this stigma. When the pressure is not adjusted in time, it will be transformed into suicidal behaviors or other mental illness behavioral characteristics. This makes LGBT victimization one of the biggest risk factors for suicide. And the biggest factor that causes LGBT victimization is bullying that almost all articles mentioned, whether it is school bullying, internet bullying or sexually-oriented bullying. Humphries’s research revealed that the students who experienced both traditional bullying and electronic bullying had higher prevalence of suicide attempt than those who experienced only one form, but the interactions for both forms showed no association, which suggest that these two forms of bullying were not synergistic in the risk of suicidality (Humphries et al., 2021). However, another study by Duong showed that both school bullying and cyber bullying were significant respectively, and suffering both together further increase the probability of suicide among LGBT teenagers. However, they also found that with the support of teachers, the association between bullying and suicide disappeared. Goodenow also reported the same conclusion that teacher support is a protective factor to prevent suicide behavior (Goodenow et al., 2006).

The safety environment of school has appeared in many studies as a strong protective factor (Duong & Bradshaw, 2014; Rimes et al., 2019; Taliaferro & Muehlenkamp, 2017). School as the first environment in which students live except for the family is very critical. If a student cannot perceive a sense of security, sympathetic and approachable in school, they will be reluctant to speak out even if they are serious bullied (McDermott et al., 2018). If they have been in such a harsh environment for a long time, they will want to resort to substance use, dropping out or even suicide attempt to escape (Goodenow et al., 2006). These conclusions all verify the stress-buffering theory (Cohen & Wills, 1985) and point to the importance of school and teacher support in suicide prevention. Ongoing professional development and in-service training for teachers should address this issue.

The anti-bullying policy has been verified by multiple studies to be an effective preventive measure against suicide attempt (Goodenow, Szalacha, & Westheimer, 2006; Hatzenbuehler & Keyes, 2013). Hatzenbuehler and Keyes’s research showed that the prevalence of suicide attempt among sexual minority students in schools that have successfully implemented this policy has dropped to 17%. Schools that have not implemented the policy have the suicide attempt rate of 31%. In addition, the research also revealed that the suicide rate of heterosexual teenagers will also be alleviated by implementing this policy. However, the policy has no significant effect on bisexual youth, indicating that the protective factors of homosexuality may be distinguished from those of bisexuality. The same policy cannot be extended to all sexual minorities.

### Strength

At our knowledge, this review is the first to focus on risk and protective factors of suicide attempts in sexual minority youth. The main strength of this study is to summarize all associated factors on suicide attempt in LGBT adolescents and contextualized into 3 different categories, finally classified as risk factors and protective factors. In addition, this literature review study summarizes the risk and protective factors of suicide attempt in different countries and regions through different perspectives in epidemiology, sociology, cultural, and political beliefs across multiple disciplines. These findings can provide a strong theoretical basis for subsequent policy formulation and implementation. Risk factors associated with suicide attempt in sexual minority were summarized in gender, ethnic minorities, childhood trauma, psychiatric symptoms, and addictive behaviors. Considering the higher prevalence in female, special attention is needed and different prevention strategies should be developed for gays and lesbians respectively.

### Limitation

However, this review also has several limitations. The first is that the number of studies included is small. Although lots of studies have reported on the suicide of adolescents with sexual minorities, but most of them have focused on the prevalence of suicidality rather than influencing factors. Moreover, most studies were concentrated in North America, only a few have focused on Europe, no relevant research can be found in Asia or Africa. Furthermore, some studies researched the influencing factors but did not perform multiple logistic regressions, which resulted in that it is impossible to obtain important evidence support for risk or protective factors of suicide attempt. Lastly, different studies have different definitions of associated factors, and standards cannot be unified. In our review, we collected the identified risk or protective factors of suicide attempts from studies only focus on sexual minorities. A large part of published studiers does not compare risk factors among sexual minorities and heterosexual participants but analyzed sexuality as specific risk factor. Therefore, only when the study clearly divided the study population according to sexual orientation into subgroups, we could specifically collect the significant risk factors from the sexual minority subgroups.

## Conclusion

Whereas risk factors associated with suicide attempt in general population have been found (female, ethnic minorities, trauma, psychiatric and addiction dimension), more specific risk factors related to sexuality have to be searched considering intimate and public environment. bullying, early coming out, not acceptable by families, not satisfied with LGBT friendship, too few numbers of friends. The protective factors of suicide attempt in LGBT population were feeling safety at school, teacher support, anti-bullying policy, and other adult’s support.

Although suicide risk assessment is particularly important with individuals experiencing mental illness, it should be aware that these factors according to LGBT youth are also independently associated with suicidality. Effective preventive measures for suicide attempt among LGBT youth need to be developed and implemented. Societal-level anti-stigma interventions may be required to reduce LGBT victimization. The impact of interventions targeting teacher responses to LGBT stigma, discussion of LGBT issues in class and reactions of family and friends to the coming out of LGBT youth should be further investigated.

## Data Availability

All data produced in the present work are contained in the manuscript

## Author’s contributions statement

Xavier Wang- Conceptualization, Research implementation, Data extraction, Writing-original draft.

Quan Gan- Research implementation, Data extraction, Writing-Review and Editing.

Junwen Zhou- Methodology, Writing-Review and Editing.

Mireille Cosquer- Writing-Review and Editing.

Bruno Falissard- Methodology, Writing-Review and Editing.

Emmanuelle Corruble- Supervision, Writing-Review and Editing.

Catherine Jousselme- Conceptualization, Methodology, Resources, Supervision.

Florence Gressier- Conceptualization, Methodology, Supervision, Writing-Review and Editing.

All co-authors reviewed and approved the manuscript prior to submission

## Acknowledgments

We sincerely thank Helene RAFFIN, Aminata ALI, Camile PREVOST for their helpful input on this article. And we also thank David Gregory BARNS to provide proofreading for this study.

