## Appendices for "A systematic review of the factors associated with suicide attempts among sexual-minority adolescents"

Table: Quality assessment of the studies

| Author | selection | | | | comparability | | outcome | | Total |
| --- | --- | --- | --- | --- | --- | --- | --- | --- | --- |
|  | representativeness of the sample | sample size | non respondents | ascertainment of the exposure | controls for the most important factor | control for any additional factor | assessment of the outcome | statistical test |  |
| Huphries et al. 2020 | 1 | 1 | 1 | 2 | 1 | 1 | 1 | 1 | 9 |
| Busby et al. 2020 | 1 | 0 | 1 | 2 | 1 | 1 | 1 | 1 | 8 |
| Turpin et al, 2020 | 1 | 1 | 1 | 2 | 1 | 1 | 1 | 1 | 9 |
| Rimes et al, 2019 | 1 | 1 | 0 | 2 | 0 | 0 | 1 | 1 | 6 |
| Toomey et al, 2019 | 1 | 1 | 0 | 2 | 1 | 1 | 2 | 1 | 9 |
| McDermott et al, 2017 | 1 | 1 | 0 | 2 | 0 | 0 | 0 | 1 | 4 |
| Taliaferro et al, 2017 | 1 | 1 | 1 | 2 | 1 | 1 | 1 | 1 | 9 |
| Duong et al, 2014 | 1 | 1 | 1 | 2 | 1 | 1 | 1 | 1 | 9 |
| Hatzenbuehler et al, 2013 | 1 | 1 | 1 | 1 | 1 | 0 | 2 | 1 | 8 |
| Mustanski et al, 2013 | 1 | 1 | 1 | 2 | 1 | 1 | 2 | 1 | 10 |
| Goodenow et al, 2006 | 1 | 1 | 1 | 2 | 1 | 1 | 2 | 1 | 10 |
| Heerngen et al, 2000 | 1 | 1 | 0 | 2 | 1 | 1 | 2 | 1 | 9 |
